# Multimorbidity and risk of incident dementia: role of disease clusters and genetic risk for dementia in a cohort of 206,960 participants

**DOI:** 10.1101/2022.07.06.22277310

**Authors:** Catherine M. Calvin, Megan C. Conroy, Sarah F. Moore, Elżbieta Kuźma, Thomas J. Littlejohns

**Affiliations:** Nuffield Department of Population Health, University of Oxford, Oxford, UK, OX3 7LF; College of Medicine and Health, University of Exeter, Exeter, UK, EX1 2LT; Albertinen-Haus Centre for Geriatrics and Gerontology, University of Hamburg, Hamburg, Germany

## Abstract

**Importance:** Individual conditions have been identified as risk factors for dementia, however, it is important to consider the role of multimorbidity as conditions often co-occur.

**Objective:** To investigate whether multimorbidity is associated with incident dementia, and whether associations vary by different clusters of disease, and genetic risk for dementia.

**Design:** A population-based prospective study.

**Setting:** The UK Biobank cohort.

**Participants:** 206,960 dementia-free women and men aged ≥60 years old at baseline Exposures: Medical conditions were captured as part of a nurse-led verbal interview conducted at assessment centres. The presence of ≥2 long-term conditions from a preselected list of 42 conditions was used to define multimorbidity. High genetic risk for dementia was based on presence of one or two Apolipoprotein (APOE) ε4 alleles.

**Main outcome:** Incident dementia was derived from hospital inpatient and death registry records.

**Results:** 89,201 (43%) participants had multimorbidity. Over a mean of 11.8 years (standard deviation=2.2), 6,182 participants developed dementia. The incidence rate per 1,000 person years was 1.87 (95% Confidence Interval [CI] 1.80-1.94) and 3.41 (95% CI 3.30-3.53) for those without and with multimorbidity, respectively. In Cox-proportional-hazards models adjusted for age, sex, ethnicity, education, socioeconomic status and APOE-ε4 carrier status, multimorbidity was associated with a 63% increased risk of incident dementia (Hazard Ratio [HR]=1.63, 95% CI 1.55-1.71). The highest dementia risk was observed for the hypertension/diabetes/coronary heart disease (HR=2.20, 95% CI 1.98-2.46) and pain/osteoporosis/dyspepsia (HR=2.00, 95% CI 1.68-2.37) clusters in females and diabetes/hypertension (HR=2.24, 95% CI 1.97-2.55) and coronary heart disease/hypertension/stroke clusters (HR=1.94, 95% CI 1.71-2.20) in males, compared to no multimorbidity. The relative associations were stronger in those with a lower genetic risk of dementia, but the absolute difference in risk between absence and presence of multimorbidity was greater in those with a higher genetic risk for dementia.

**Conclusions and Relevance:** Multimorbidity was strongly associated with an increased risk of dementia. The strength of associations varied by clusters of disease and genetic risk for dementia. These findings could help with the identification of individuals at high risk of dementia as well as the development of targeted interventions to reduce or delay dementia incidence.

## Introduction

Due to increasing life expectancy the global prevalence of dementia is projected to triple within 30 years from 57 to 153 million by 2050^1^. Despite the increasing incidence there is evidence that the age-specific prevalence of dementia is declining^2^. This decline in prevalence could partially be due to improvements in the control and treatment of cardiovascular and metabolic diseases, such as hypertension, stroke and diabetes ^3^. These, and other health-related conditions, have previously been identified as potentially key modifiable risk factors for dementia reduction^4,5^.

However, conditions often do not occur in isolation, with approximately a third of the global population living with two or more long-term health conditions, termed multimorbidity^6,7^. Similar to dementia, the prevalence of multimorbidity increases substantially with age, with two thirds of 65-84 year olds and more than 80% of those aged 85 years or older living with multiple conditions^8^. Consequently, when investigating the role of certain health conditions as risk factors for dementia it is important to understand the role of multiple conditions, in particular whether certain clusters of disease are differentially associated with dementia risk^9^.

To our knowledge, two studies have investigated the association between multimorbidity and dementia risk^10,11^. Both found that multimorbidity was associated with an increased risk of dementia, whilst one found that neuropsychiatric, cardiovascular and sensory impairment/cancer clusters but not a respiratory/metabolic/musculoskeletal cluster were associated with an increased risk^11^. The same study found no evidence that Apolipoprotein (APOE) ε4, the strongest genetic risk factor for dementia^12^, modified these associations^11^. However, these studies consisted of between 500 and 700 incident dementia cases and might have lacked sufficient statistical power to detect whether certain disease clusters are associated with different risks of dementia and whether genetic risk for dementia modifies these associations^10,11^.

In the current study of more than 200,000 participants and 6,200 incident dementia cases we investigated 1) whether multimorbidity and 2) multimorbidity clusters were associated with incident dementia, and 3) whether any observed associations were modified by genetic risk for dementia.

## Methods

### Population

Between 2006-2010, half a million women and men aged between 40 and 69 years old joined the UK Biobank study^13,14^. All participants attended one of 22 baseline assessment centres located throughout England, Scotland and Wales and provided electronic-signed consent. At assessment, participants provided sociodemographic, lifestyle and health-related information through a touchscreen questionnaire and nurse-led verbal interview, and underwent a range of physical examinations. Blood samples were obtained and used to perform genome-wide genotyping at a later date. UK Biobank received ethical approval from the National Health Service North West Centre for Research Ethics Committee (Ref: 11/NW/ 0382). To restrict to individuals at risk of developing dementia over the follow-up period we excluded participants aged <60 years at baseline.

### Multimorbidity

Participants self-reported medical conditions during the nurse-led verbal interview. The interviewer was guided by a tree structure based on the International Classification of Disease (ICD) coding system in order to ensure entries were standardised^15^. In the current study, the diseases included in the multimorbidity definition were based on the list developed by Barnett and colleagues^8^ (see **eTable 1** for list of included conditions)^16^. Multimorbidity was defined as the presence of at least two of the 42 conditions. Participants with zero or one condition were defined as not having multimorbidity and formed the reference group in the analyses.

### Dementia

Dementia was ascertained using hospital inpatient and death registry records. Primary and secondary hospital diagnoses and causes of deaths were recorded using the ICD coding system. The ICD codes for dementia were previously selected and validated by the UK Biobank outcome adjudication group and are listed in **eTable 2**^17^. Participants with a diagnosis of dementia prior to study baseline, or who self-reported having dementia during the verbal interview at baseline assessment, were excluded from the current study.

### Covariates

Sociodemographic covariates included age in years, sex (‘women’, ‘men’), ethnicity (‘white’, ‘non-white’), education (‘college, university or professional qualification’, ‘secondary school or vocational qualification’, ‘no qualification’), tertiles of socioeconomic status measured with the Townsend deprivation score combining information on social class, employment, car availability, and housing ^18^. Apolipoprotein E (APOE)-ε4 carrier status (‘non-carrier’, ‘carrier’) was determined using the rs429358 and rs7412 single nucleotide polymorphisms, which were directly genotyped on the UK Biobank arrays ^19^.

### Statistical analysis

#### 1) All analyses

Cox proportional-hazards models adjusted for age, sex, ethnicity, education, socioeconomic status and APOE-ε4 carrier status were used to estimate the association between multimorbidity presence and clusters and incident dementia. Person-years were calculated from date of attending baseline assessment until date of first dementia diagnosis, date of death, date lost to follow-up or end of follow-up, whichever occurred first. End of follow-up was based on the availability of the medical record data in UK Biobank, which was censored at 30^th^ September 2021 for England, 31^st^ July 2021 for Scotland, and 28^th^ February 2018 for Wales. All models were assessed for the proportionality of hazards assumption using log-log plots, Kaplan-Meier observed survival curves, and post-hoc tests of proportional hazards with Schoenfield residuals. Sex violated the proportional hazards assumption and was entered into the models in strata. Participants with missing data or answered ‘prefer not to answer/do not know’ constituted <5% of the sample and were excluded from the analyses. Dementia incidence rates (IR) per 1,000 person years were calculated for each exposure group.

P-values were 2-sided, and the type I error rate for statistical significance was set at .05. Analyses were performed using Stata SE version 17.0 (StataCorp LLC, College Station, TX). RStudio 1.4.1717 (*poLCA* package) was used to derive multimorbidity clusters for the latent class analysis.

#### 2) Presence of Multimorbidity

In the main analysis we investigated the association between multimorbidity (≥2 conditions) compared to no multimorbidity (≤1 condition) and incident dementia. In a sensitivity analysis, we repeated the main analysis using three separate follow-up periods of ≤5 years, >5 to ≤10 years and >10 years. This was to explore the potential effect of reverse causation due to dementia’s long prodromal period whereby the underlying pathology of preclinical dementia may affect health several years prior to a clinical diagnosis ^21^. In secondary analyses, we investigated the possibility of a dose-response relationship by categorizing multimorbidity as ≤1 condition, 2, 3, 4, 5 and ≥6 conditions. To investigate potential effect modification by sociodemographic factors, interaction terms were first entered into the main model for multimorbidity by 1) age (<65, ≥65), 2) sex, 3) education, 4) socioeconomic status, and then stratified analyses were performed within each group.

#### 3) Multimorbidity clusters

Latent class analysis was used to determine multimorbidity clusters, allocating each participant with multimorbidity to a single non-overlapping cluster while allowing health conditions to contribute by varying probabilities to multiple clusters^22^. Clusters were estimated separately for men and women, as output from exploratory models of the total sample showing several sex-dominant cluster groups (see **eTable 3**). A random *training* sample of 80% of participants with multimorbidity was used to determine the optimal number of clusters in men and women, and subsequently to estimate the effect of multimorbidity clusters on dementia risk. Output statistics were generated for multiple latent class analysis models of between 1 to 12 cluster solutions, and the optimal number of clusters were determined using a combination of sample-size adjusted Bayesian Information Criteria statistics, clinical judgement, and capping the smallest cluster to >5% of the training sample. Each cluster within men and women was characterised by the three health conditions with the highest probabilities above 5% of contributing to that cluster, excluding conditions where their observed prevalence (O) was equal to or less than that of the total population of men and women: expected prevalence (E).

To assess the validity of the determined cluster solutions, conditions from the remaining 20% of men and women with multimorbidity (a *test* sample) were entered into latent class analysis models, setting the number of clusters to match the optimal number from the training set. The characterisation and relative size of the clusters determined from the training and test samples were compared, as were their associations with dementia risk in Cox proportional-hazards models.

#### 4) Effect modification by APOE-ε4

An interaction term for multimorbidity and APOE-ε4 carrier status was first entered into the main models and then stratified analyses were performed within non-APOE-ε4 and APOE-ε4 carrier subgroups. These analyses were repeated for each multimorbidity cluster.

## Results

Of 502,412 participants, 284,943 aged less than 60 at baseline, 166 with prevalent dementia and 10,343 with missing covariate information were excluded, resulting in a final sample of 206,960 participants. Of these, 89,201 (43%) had multimorbidity at baseline. Participants with multimorbidity were more likely to be older, women, of non-white ethnicity, have lower educational qualifications, and be from more socioeconomically deprived areas compared to those with no multimorbidity (**Table 1**). No difference in APOE-ε4 carrier status was observed by presence or absence of multimorbidity. See **eTable 4** for baseline characteristics by incident dementia. A total of 6,182 (3%) participants developed dementia over 2,451,957 person-years of follow-up (mean=11.8, standard deviation=2.2).

**Table 1.**
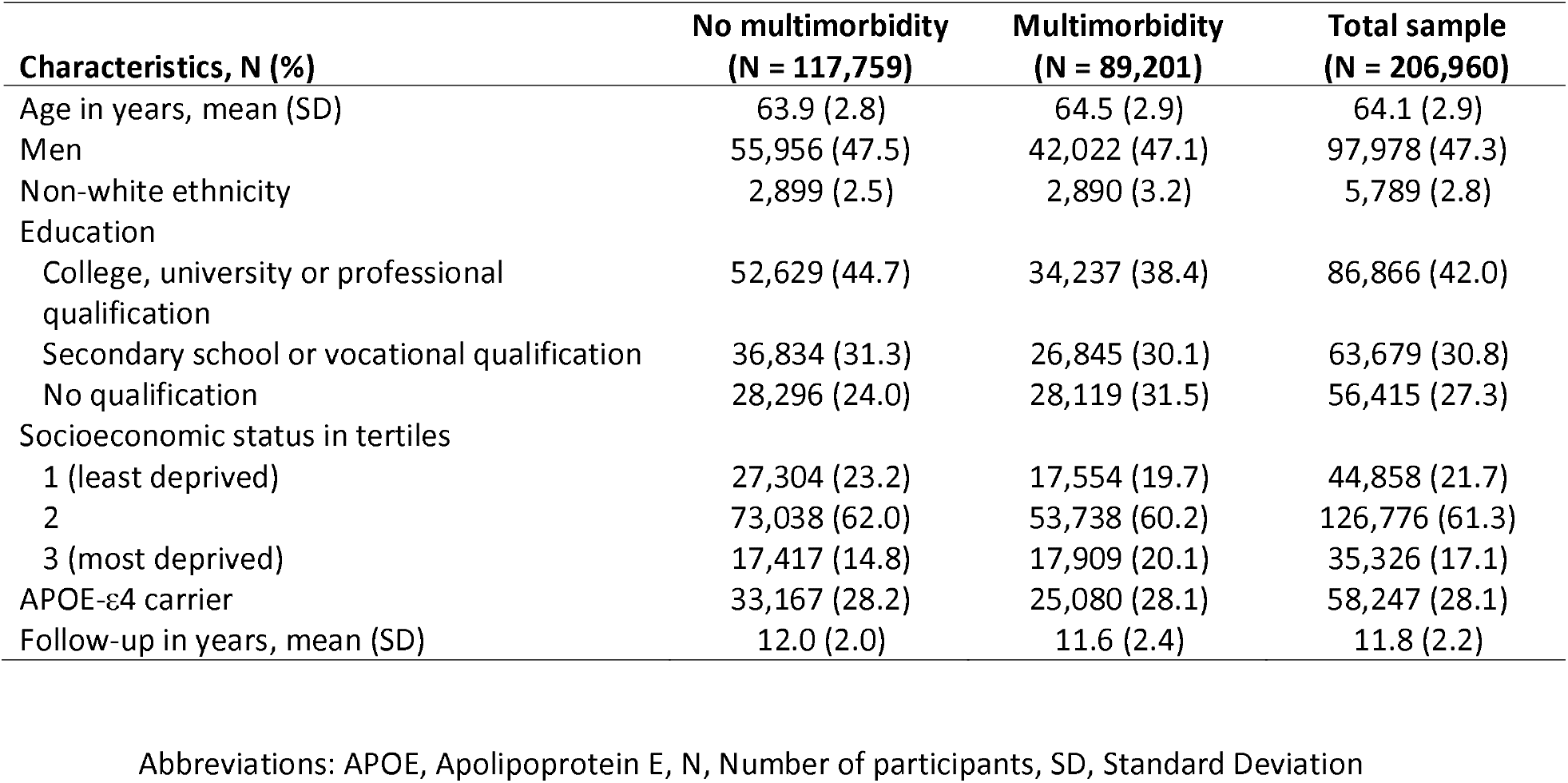
Baseline characteristics by multimorbidity

### 1) Multimorbidity

The IR of dementia was almost double in participants with multimorbidity compared to no multimorbidity (**Figure 1**), IR = 3.41 (95% Confidence Interval [CI] 3.30-3.53) vs IR=1.87 (95% CI 1.80-1.94) per 1,000 person-years, respectively. In fully adjusted Cox proportional-hazards models, multimorbidity was associated with a 63% increased risk of dementia compared to no multimorbidity (Hazard Ratio [HR] = 1.63, 95% CI 1.55-1.71; **Figure 1**). The pattern of associations remained similar when restricting to different periods of follow-up (**Figure 1**). For instance, for participants with ≥10 years of follow-up, the HR for multimorbidity in association with incident dementia was 1.59 (95% CI 1.47-1.71). When investigating number of conditions with dementia risk, a strong dose-response association was observed (**Figure 2**). The HRs were 1.41 (95% CI 1.32-1.50), 1.61 (95% CI 1.49-1.73), 2.19 (95% CI 1.99-2.40), 2.59 (95% CI 2.26-2.96) and 3.15 (95% CI 2.65-3.75) for 2, 3, 4, 5 and ≥6 conditions, respectively, compared to no multimorbidity.

**Figure 1.**
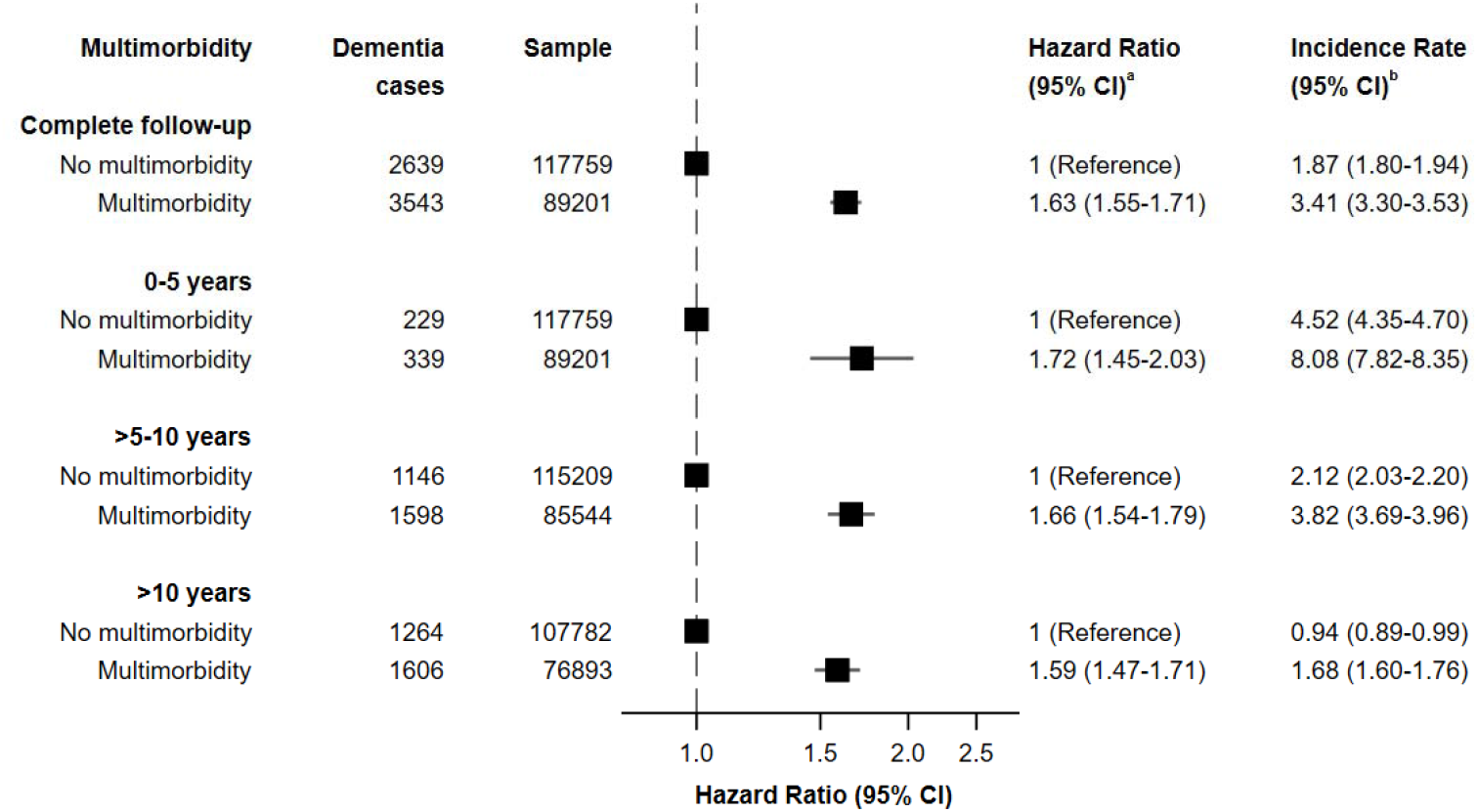
Cox-proportional hazards models for the association between multimorbidity and incident dementia by different follow-up periods Abbreviations: CI, Confidence Interval. ^a^All models adjusted for age, ethnicity, education, socioeconomic status and APOE-ε4 ^b^Incidence rate per 1,000 person-years

**Figure 2.**
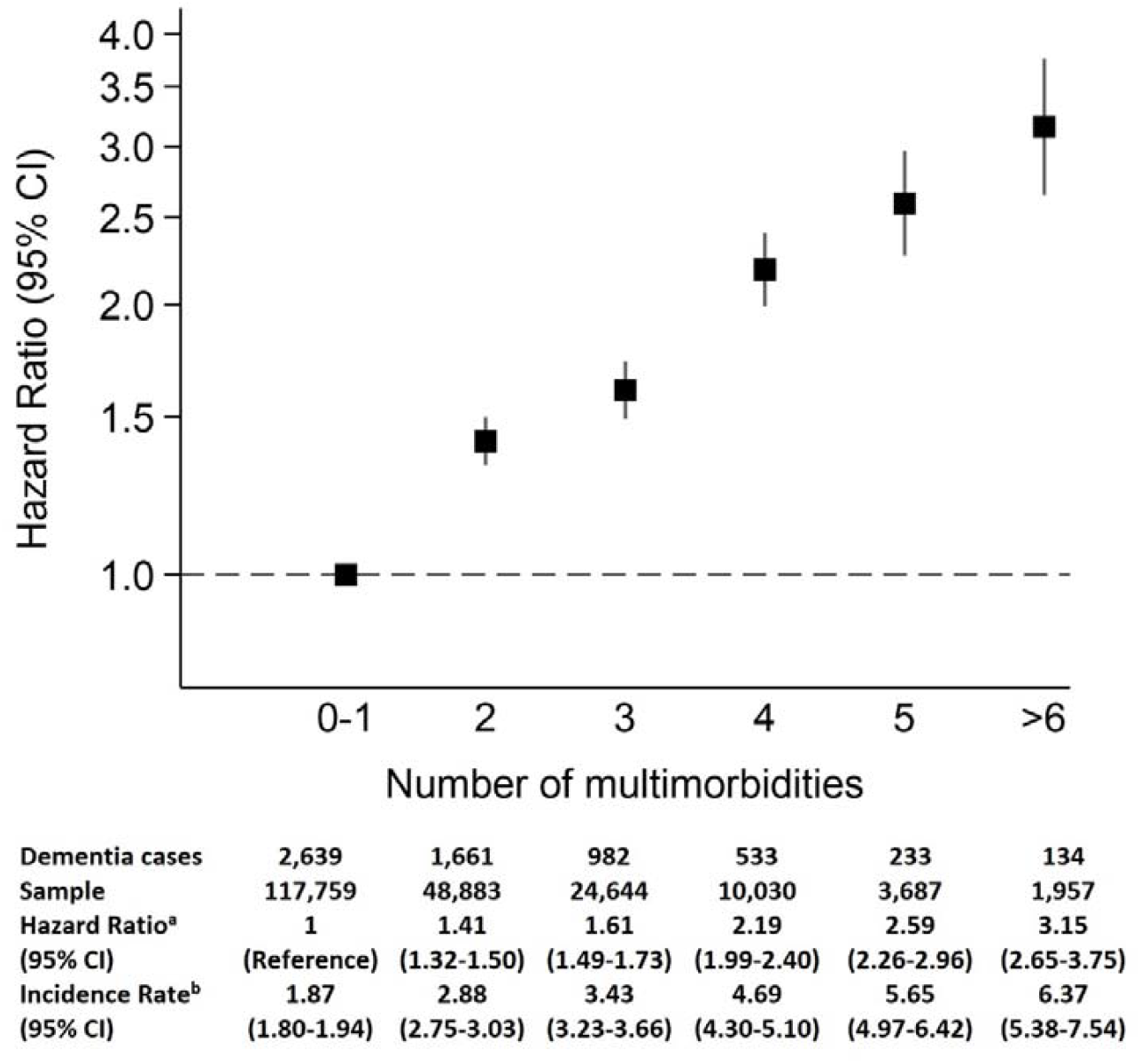
Cox-proportional hazards models for the association between number of multimorbid conditions and incident dementia Abbreviations: CI, Confidence Interval. ^a^All models adjusted for age, ethnicity, education, socioeconomic status and APOE-ε4 ^b^Incidence rate per 1,000 person-years

There was no evidence for a statistically significant interaction between multimorbidity and age (*p-value*=0.10), sex (*p-value*=0.36), education (*p-value*=0.94) or socioeconomic status (*p-value*=0.24) (**eTable 5**).

### 2) Multimorbidity clusters

41 conditions were included in the analysis for women (prostate disorders excluded) and 40 for men (polycystic ovaries and endometriosis excluded). In a comparison of latent class analysis models, seven clusters were identified as the optimal number for women and six for men (**eFigure 1a** and **1b**). A thyroid disease cluster was unique to women, and a coronary heart disease (CHD)/diabetes cluster was unique for men (see **eTable 6 and 7**). There was high similarity in the characteristics of the cluster groups in the training samples versus those of the test samples for men and women respectively (see **eTable 8**). All clusters of multimorbidity in males and females were associated with an increased risk of dementia compared to no multimorbidity (**Table 2**). In females, the hypertension/diabetes/CHD (HR = 2.20, 95% CI 1.98-2.46) and pain/osteoporosis/dyspepsia (HR=2.00, 95% CI 1.68-2.37) clusters, and in males, the diabetes/hypertension (HR=2.24, 95% CI 1.97-2.55) and CHD/hypertension/stroke (HR=1.94, 95% CI 1.71-2.20) clusters, were associated with double of more the risk of incident dementia compared to no multimorbidity.

**Table 2.** Sex stratified Cox-proportional hazards models for the association between multimorbidity clusters and incident dementia

| Multimorbidity clusters | Dementia cases | Sample | Median morbidities [Q1-Q3] | Hazard Ratio (95% CI)<sup>a</sup> | Incident Rate (95% CI)<sup>b</sup> |
| --- | --- | --- | --- | --- | --- |
| **Female** |  |  |  |  |  |
| No multimorbidity | 1,231 | 61,803 | 1 [0–1] | 1 (reference) | 1.64 (1.55, 1.74) |
| Hypertension (100%), diabetes (23%), CHD (15%) | 452 | 9,107 | 2 [2-3] | 2.20 (1.98-2.46) | 4.22 (3.85, 4.63) |
| Cancer (100%) | 188 | 6,440 | 2 [2-3] | 1.37 (1.17-1.60) | 2.52 (2.19, 2.91) |
| Thyroid disorders (100%) | 173 | 5,387 | 3 [2-3] | 1.44 (1.23-1.69) | 2.68 (2.31, 3.11) |
| Pain (63%), dyspepsia (37%), depression (23%) | 155 | 5,184 | 2 [2-4] | 1.43 (1.21-1.69) | 2.47 (2.11, 2.89) |
| Asthma (100%), COPD (12%) | 161 | 4,742 | 3 [2-4] | 1.66 (1.41-1.95) | 2.86 (2.45, 3.34) |
| Pain (100%), hypertension (100%) | 120 | 3,578 | 2 [2-3] | 1.42 (1.18-1.72) | 2.79 (2.33, 3.33) |
| Pain (46%), osteoporosis (25%), dyspepsia (24%) | 146 | 3,307 | 2 [2-3] | 2.00 (1.68-2.37) | 3.74 (3.18, 4.40) |
| **Male** |  |  |  |  |  |
| No multimorbidity | 1,408 | 55,956 | 1 [0-1] | 1 (reference) | 2.12 (2.01, 2.23) |
| Hypertension (100%), pain (43%), dyspepsia (19%) | 349 | 9,259 | 2 [2-3] | 1.38 (1.22-1.55) | 3.24 (2.92, 3.60) |
| CHD (100%), hypertension (74%), stroke (9%) | 313 | 5,603 | 3 [2-4] | 1.94 (1.71-2.20) | 5.02 (4.49, 5.61) |
| Asthma (100%), COPD (13%), psoriasis (9%) | 173 | 5,051 | 3 [2-3] | 1.32 (1.12-1.54) | 2.97 (2.56, 3.45) |
| Pain (54%), dyspepsia (34%), prostate disorders (24%) | 197 | 4,846 | 2 [2-3] | 1.52 (1.31-1.77) | 3.47 (3.02, 4.00) |
| Diabetes (100%), hypertension (85%) | 281 | 4,742 | 3 [2-3] | 2.24 (1.97-2.55) | 5.28 (4.70, 5.94) |
| Cancer (100%) | 149 | 4,088 | 2 [2-3] | 1.41 (1.19-1.66) | 3.35 (2.85, 3.93) |

Abbreviations: CI, Confidence Interval, CHD, Coronary Heart Disease, COPD, Chronic Obstructive Pulmonary Disease, Q, Quartile
<sup>a</sup> All models adjusted for age, ethnicity, education, socioeconomic status and APOE-ε4
<sup>b</sup> Incidence rate per 1,000 person-years

### 3) Effect modification by APOE-ε4

Multimorbidity significantly interacted with APOE-ε4 carrier status (*p-value*<.0001) in association with incident dementia. When stratifying by APOE-ε4 carrier status, the association between multimorbidity and incident dementia was stronger in non-APOE-ε4 carriers (HR=1.96, 95% CI 1.81-2.11) and attenuated in APOE-ε4 carriers (HR=1.39, 95% CI 1.30-1.49). Similarly the associations were generally stronger within non-APOE-ε4 carriers for multimorbidity clusters for both women (**Figure 3a**) and men (**Figure 3b**). However, despite stronger relative risks in non-APOE-ε4 carriers, the absolute risks are higher in APOE-ε4 carriers. For example, among non-APOE-ε4 carriers the IRs were 2.33 (95% CI 2.23-2.45) and 1.06 (95% CI 1.00-1.12) per 1,000 person years for those with and without multimorbidity, respectively. Whilst, among APOE-ε4 carriers the IRs were 3.92 (95% CI 3.73-4.13) and 6.12 (95% CI 5.83-6.41) per 1,000 person years for those with and without multimorbidity, respectively. Therefore, the risk difference, calculated by subtracting the no multimorbidity IRs from the multimorbidity IRs, were higher within APOE-ε4 carriers: 2.20 per 1,000 person years in APOE-ε4 carriers vs 1.27 per 1,000 person-years in non-APOE-ε4 carriers. Similar findings were observed for multimorbidity clusters (**Figure 3a** and **3b**).

**Figure 3.**
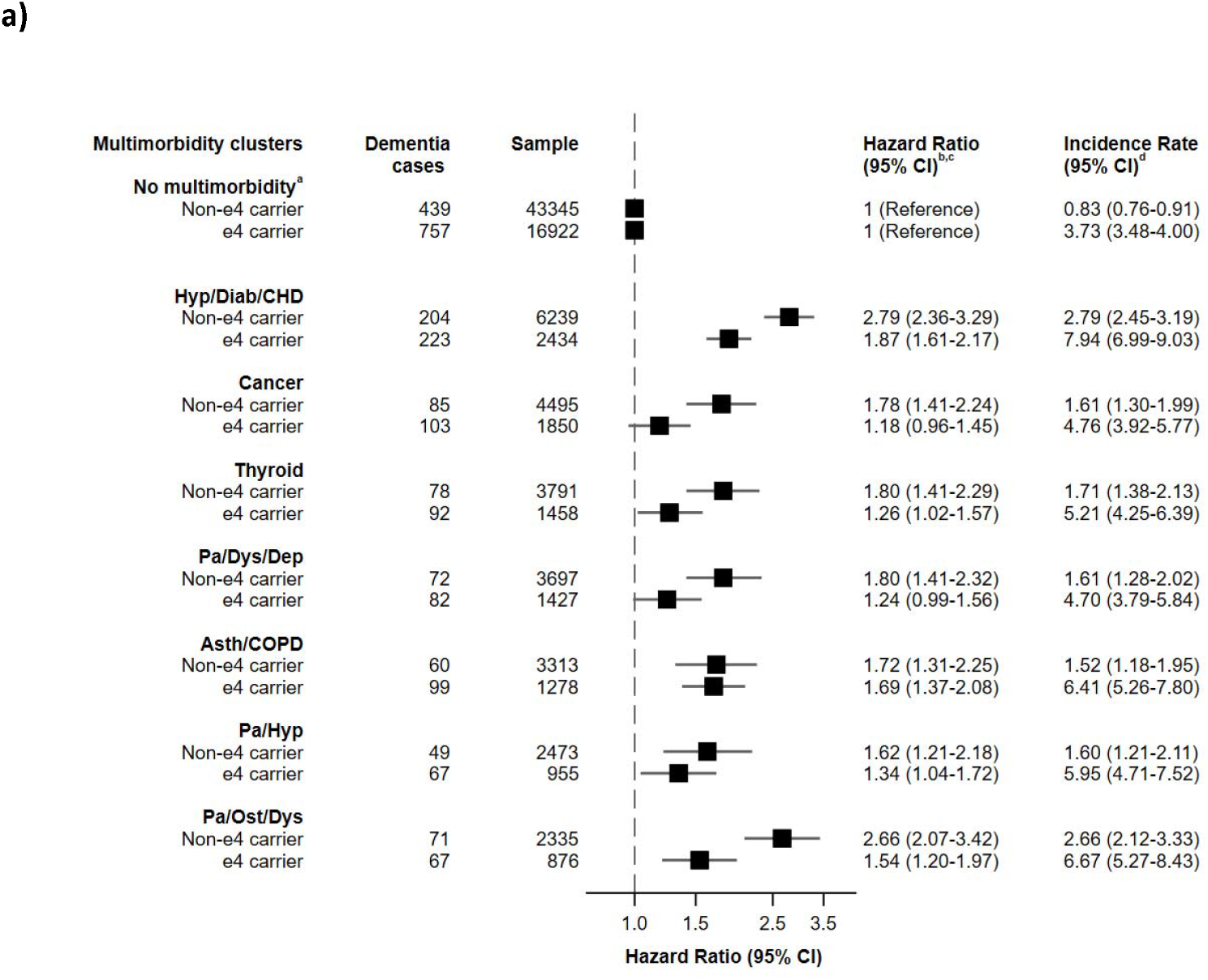
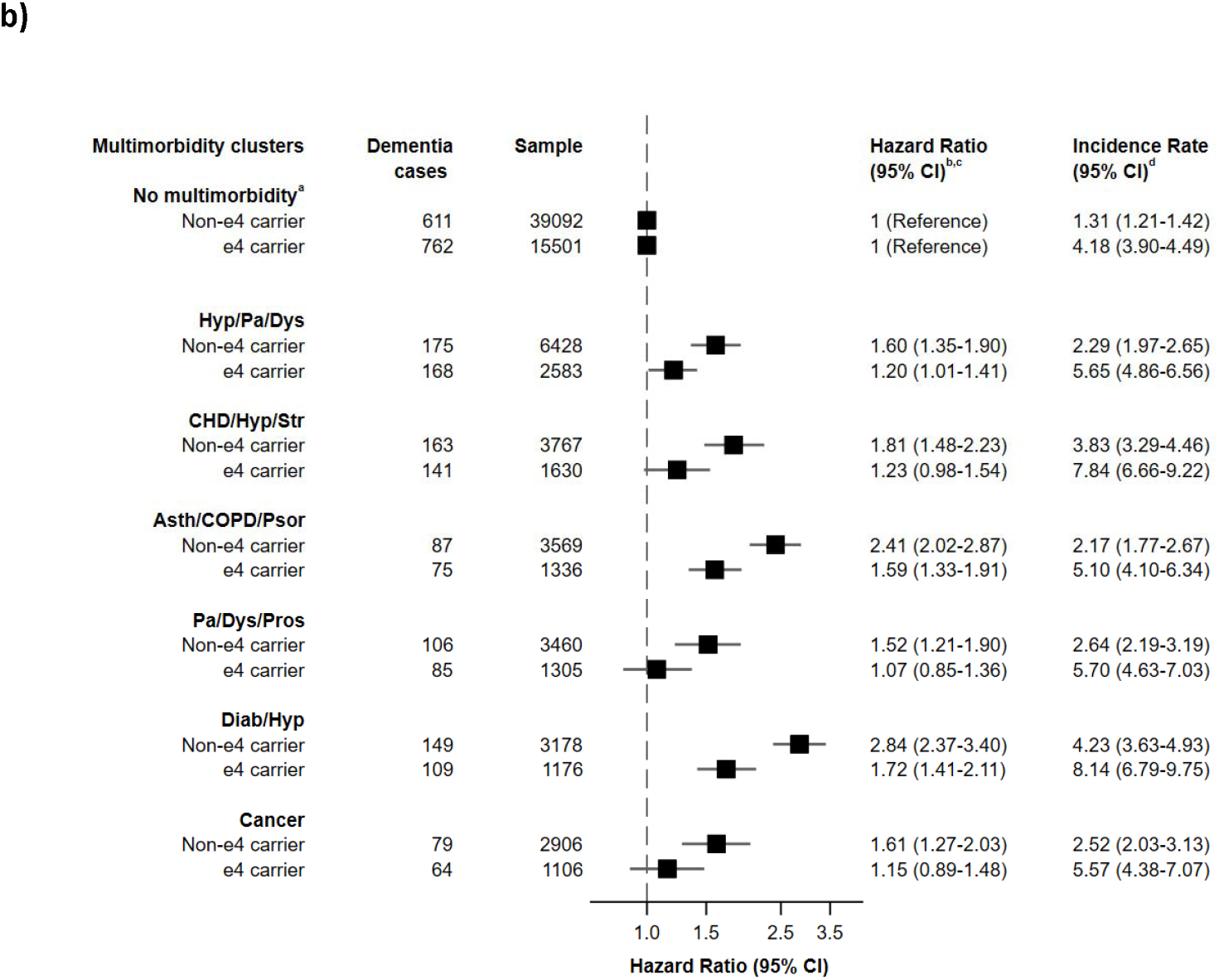
Cox-proportional hazards models of the interaction between multimorbidity clusters and APOE and incident dementia in **a)** women and **b)** men Abbreviations: Asth, Asthma, CI, Confidence Interval, CHD, Coronary Heart Disease, COPD, Chronic Obstructive Pulmonary Disease, Dep, Depression, Diab, Diabetes, Dys, Dyspepsia, Hyp, Hypertension, Ost, Osteoporosis, Pa, Pros, Prostate Disorders, Pain, Psor, Psoriasis, Str, Stroke. ^a^No multimorbidity non-ε4 carrier serves as reference group for non-ε4 clusters and ε4 carrier serves as reference group for ε4 carrier clusters ^b^All models adjusted for age, ethnicity, education, socioeconomic status and APOE-ε4 ^c^P-value for interaction for each multimorbidity cluster by APOE status are as follows: Women, Hyp/Diab/CHD *p*=<0.001, Cancer *p*=0.01, Thyroid *p*=0.03, Pa/Dys/Dep *p*=0.03, Asth/COPD *p*=0.91, Pa/Hyp *p*=0.33 and Pa/Ost/D *p*=0.003. Men, Hyp/Pa/Dys *p*=0.02, CHD/Hyp/Str *p*=0.01, Asth/COPD/Psor *p*=0.001, Pa/Dys/Pros *p*=0.04, Diab/Hyp *p*=<0.001, Cancer p=0.06. ^d^Incidence rate per 1,000 person-years

## Discussion

In this large UK-based cohort of more than 200,000 participants aged 60 years or older, presence of multimorbidity was associated with a 63% increased risk of developing dementia over 15 years of follow-up. A strong dose-response association was observed between number of conditions and dementia risk. Various multimorbidity clusters were differentially associated with dementia risk, with hypertension/diabetes/CHD and pain/osteoporosis/dyspepsia clusters in females and diabetes/hypertension and CHD/hypertension/stroke clusters in males associated with double or more the risk of dementia. The associations were consistently stronger amongst individuals with a lower genetic risk for dementia based on APOE-ε4 carrier status. However, the risk differences between absence and presence of multimorbidity were larger in those with a higher genetic risk for dementia.

Our findings are consistent with a recent study of 10,095 UK-based adults followed over 32 years^10^. Ben Hassen and colleagues found that multimorbidity was associated with an increased risk of dementia and that there was a dose-response association between number of conditions and dementia risk^10^. However, to our knowledge, only one previous study has explored multimorbidity and different clusters of disease in association with dementia risk. In 2,478 Swedish-based adults, Grande and colleagues found that neuropsychiatric, cardiovascular, and sensory impairment/cancer, but not respiratory/metabolic/musculoskeletal clusters of multimorbidity were associated with an increased risk of dementia over 12 years follow-up^11^. We identified similar clusters in association with dementia, however, we also found that clusters driven by respiratory and metabolic conditions were associated with dementia risk. Grande and colleagues included 506 dementia cases compared to the 6,182 cases included in the current study, so they might have lacked the statistical power to detect certain disease clusters in association with dementia.

Grande and colleagues also found that the associations between cardiovascular and neuropsychiatric clusters and dementia risk was stronger amongst APOE-ε4 carriers, although the test for interaction was not statistically significant^11^. In contrast, we found a statistically significant interaction between APOE genotype and multimorbidity and dementia risk, finding stronger associations in non-APOE-ε4 carriers. The pattern of associations remained similar for multimorbidity clusters. Presence of the APOE-ε4 allele is responsible for around a quarter of heritability for the most common form of dementia, Alzheimer’s disease^23^. APOE-ε4 has been linked with an increased risk of various vascular diseases^24,25^, with inconsistent associations observed for non-vascular diseases, such as depression^26^ and cancer^27^.

Our observation of a stronger association in non-APOE-ε4 carriers could be due to the substantially elevated baseline risk of dementia in APOE-ε4 carriers which could attenuate the relative risk between multimorbidity and dementia within this group. This is supported by our finding that the absolute risk differences between no multimorbidity and multimorbidity presence/clusters were greater in those with a higher genetic risk of dementia. Nevertheless, this finding could have important implications for recruitment into prevention trials for Alzheimer’s disease which are increasingly incorporating presence of APOE-ε4 alleles into the inclusion criteria^28^. If targeting disease clusters for dementia risk reduction then the inclusion of non-APOE-ε4 carriers might be recommended. In the current study the strongest associations with dementia for both sexes were identified for clusters largely driven by cardiometabolic and cardiovascular diseases, such as hypertension, diabetes, coronary heart disease and stroke. This is consistent with the recommendations from expert panels which suggest targeting these conditions for dementia risk reduction^4,5^. Such recommendations typically focus on these conditions individually, whereas our findings suggest co-occurrence of these conditions might be especially important.

We also identified clusters in women and men for less well-established risk factors, such as respiratory diseases and cancer. Meta-analyses have found that reduced pulmonary function and respiratory disease is associated with dementia incidence and death^29,30^, with reduced oxygen to the brain resulting in neuronal death as a potential causal mechanism^31^. Conversely, meta-analyses have largely found that cancer is inversely associated with dementia risk^32,33^. However, previous findings are based on cancer alone, whereas we found that cancer in the presence of other diseases was associated with an increased risk of dementia. In the current study, other clusters grouped diseases from multiple organ systems, such as pain/osteoporosis/dyspepsia in women and pain/dyspepsia/prostate disorders in men. These findings can be viewed as hypothesis generating, with future studies focusing on whether such clusters of disease could provide new insights into the risk of developing dementia.

This study has several strengths, including a large sample size, long follow-up period, and the availability of detailed and diverse data which enabled both a comprehensive definition of multimorbidity and exploration of genetic interactions. However, this study also has several limitations. The conditions used to define multimorbidity were self-reported. Incident dementia was ascertained using hospital inpatient and death registry records. Whilst studies have found these to be accurate sources of dementia diagnoses, they will underestimate cases captured in other sources such as primary care or memory clinics ^17,34^. However, misclassification errors or underreporting are likely to have biased the results toward the null. Hospital inpatient and death records also perform poorly for determining dementia subtypes, so future studies are necessary to investigate whether multimorbidity clusters vary in association with Alzheimer’s disease, vascular, mixed or other types of dementia. Due to the observational nature of the study causality cannot be inferred and residual confounding remains.

In conclusion, we found that multimorbidity was associated with increased risk of incident dementia. Dementia risk was highest in hypertension/diabetes/CHD and pain/osteoporosis/dyspepsia clusters in females and diabetes/hypertension and CHD/hypertension/stroke clusters in males. The relative risk was stronger in those with a lower genetic risk of dementia. However, the absolute difference in risk between those with and without multimorbidity was greater amongst individuals with a higher genetic risk of dementia. Overall, these findings could improve identification of individuals at high-risk of dementia and highlight the necessity of targeting clusters of diseases for dementia prevention rather than individual risk factors.

## Data Availability

All data are available upon approval by UK Biobank

https://www.ukbiobank.ac.uk/

## Author contributions

CMC, EK, and TJL conceived and designed the study. All authors were involved in the interpretation of data, drafting of the work, provided final approval of the version to be published and agree to be accountable for all aspects of the work in ensuring that questions related to the accuracy or integrity of any part of the work are appropriately investigated and resolved.

## Additional Information

This research has been conducted using the UK Biobank Resource under Application Number 41115. We are grateful to the participants for dedicating a substantial amount of time to take part in the UK Biobank study as well as the staff who make it possible.

