## Supplemental Material for "Multimorbidity and risk of incident dementia: role of disease clusters and genetic risk for dementia in a cohort of 206,960 participants"

**eTable 1 - List of 42 conditions used to define multimorbidity and their prevalence in the analytic sample**

| **Condition** | **N (%)** |
| --- | --- |
| hypertension | 74,932 (36.2) |
| painful condition | 44,420 (21.5) |
| cancer (any) | 22,143 (10.7) |
| asthma | 22,095 (10.7) |
| treated dyspepsia | 20,932 (10.1) |
| coronary heart disease | 15,691 (7.6) |
| thyroid disorders | 14,784 (7.1) |
| diabetes | 14,382 (7.0) |
| depression | 9,707 (4.7) |
| psoriasis or eczema | 6,614 (3.2) |
| prostate disorders | 6,456 (3.1) |
| rheumatoid arthritis, other inflammatory polyarthropathies & systematic connective tissue disorders | 5,667 (2.7) |
| stroke and TIA | 5,573 (2.7) |
| osteoporosis | 5,502 (2.7) |
| chronic obstructive pulmonary disease | 5,037 (2.4) |
| migraine | 4,926 (2.4) |
| irritable bowel syndrome | 4,576 (2.2) |
| glaucoma | 3,646 (1.8) |
| diverticular disease of intestine | 3,512 (1.7) |
| anxiety & other neurotic, stress related & somatoform disorders | 3,454 (1.7) |
| atrial fibrillation | 2,591 (1.3) |
| inflammatory bowel disease | 1,841 (0.9) |
| epilepsy | 1,524 (0.7) |
| chronic sinusitis | 1,314 (0.6) |
| endometriosis | 1,162 (0.6) |
| pernicious anaemia | 772 (0.4) |
| Meniere’s disease | 752 (0.4) |
| bronchiectasis | 745 (0.4) |
| chronic fatigue syndrome | 706 (0.3) |
| peripheral vascular disease | 655 (0.3) |
| schizophrenia (and related non-organic psychosis) or bipolar disorder | 633 (0.3) |
| Parkinson's disease | 623 (0.3) |
| chronic kidney disease | 607 (0.3) |
| multiple sclerosis | 577 (0.3) |
| viral hepatitis | 485 (0.2) |
| chronic liver disease | 441 (0.2) |
| heart failure | 432 (0.2) |
| alcohol problems | 245 (0.1) |
| treated constipation | 228 (0.1) |
| polycystic ovary | 61 (0.03) |
| anorexia or bulimia | 56 (0.03) |
| other psychoactive substance misuse | 19 (0.01) |

**eTable 2 - ICD codes used to ascertain dementia**

| **ICD-9** | **ICD-10** |
| --- | --- |
| 331.0, 290.4, 331.1, 290.2, 290.3, 291.2, 294.1, 331.2, 331.5 | F00, F00.0, F00.1, F00.2, F00.9, G30, G30.0, G30.1, G30.8, G30.9, F01, F01.0, F01.1, F01.2, F01.3, F01.8, F01.9, I67.3, F02.0, G31.0, A81.0, F02, F02.1, F02.2, F02.3, F02.4, F02.8, F03, F05.1, F10.6, G31.1, G31.8 |

Abbreviations: ICD, International Classification of Disease

**eTable 3 - 8-class cluster solution of multimorbidity using latent class analysis of men and women, including sex as a condition**

| Cluster^a^ | % of training sample  (n = 74,874) | Male probability | Lead condition | Subsidiary condition 1 | Subsidiary condition 2 |
| --- | --- | --- | --- | --- | --- |
| 1 | 13.1 | 0.0 | Pain (54.2%) | Dyspepsia (30.0%) | Hypertension (23.6%) |
| 2 | 15.3 | 100 | Hypertension (87.8%) | Diabetes (37.7%) | CHD (36.1%) |
| 3 | 8.8 | 100 | Pain (46.1%) | Hypertension (36.7%) | Dyspepsia (30.8%) |
| 4 | 8.1 | 7.0 | Hypertension (94.2%) | Diabetes (24.6%) | Dyspepsia (20.9%) |
| 5 | 19.8 | 40.5 | Cancer (100%) | Hypertension (60.8%) | Pain (26.7%) |
| 6 | 9.3 | 45.5 | Asthma (100%) | Hypertension (49.6%) | Pain (28.4%) |
| 7 | 18.9 | 9.5 | Thyroid (100%) | Hypertension (49.1%) | Pain (28.1%) |
| 8 | 6.9 | 48.8 | Hypertension (100%) | Pain (100%) | Dyspepsia (8.2%) |

**^a^**	In 5 out of 8 clusters there is a predominant sex, i.e. probability of men contributing to cluster is either <10% or 100%.

**eTable 4 - Baseline characteristics of participants by incident dementia**

| **Characteristics, N (%)** | **No incident dementia**  **(N = 200,778)** | **Incident dementia**  **(N = 6,182)** | **Total sample**  **(N = 206,960)** |
| --- | --- | --- | --- |
| Age in years, mean (SD) | 64.1 (2.8) | 65.7 (2.7) | 64.1 (2.8) |
| Men | 94,754 (47.2) | 3,224 (52.2) | 97,978 (47.3) |
| Non-white ethnicity | 5,583 (2.8) | 206 (3.3) | 5,789 (2.8) |
| Education |  |  |  |
| College, university or professional  qualification | 84,784 (42.2) | 2,082 (33.7) | 86,866 (41.9) |
| Secondary school or vocational qualification | 61,892 (30.8) | 1,787 (28.9) | 63,679 (30.8) |
| No qualification | 54,102 (27.0) | 2,313 (37.4) | 56,415 (27.3) |
| Socioeconomic status in tertiles |  |  |  |
| 1 (least deprived) | 43,681 (21.8) | 1,177 (19.0) | 44,858 (21.7) |
| 2 | 123,189 (61.4) | 3,587 (58.0) | 126,776 (61.3) |
| 3 (most deprived) | 33,908 (16.9) | 1,418 (22.9) | 35,326 (17.0) |
| APOE-ε4 carrier | 54,902 (27.3) | 3,345 (54.1) | 58,247 (28.1) |
| Multimorbidity | 85,658 (42.7) | 3,543 (57.3) | 89,201 (43.1) |
| Follow-up in years, mean (SD) | 11.9 (2.1) | 9.2 (2.8) | 11.8 (2.2) |

Abbreviations: APOE, Apolipoprotein E, N, Number of participants, SD, Standard Deviation.

**eTable 5 - Cox-proportional hazards models of the interaction between multimorbidity and sociodemographic characteristics and incident dementia**

|  | **No Multimorbidity** | | | | **Multimorbidity** | | | |
| --- | --- | --- | --- | --- | --- | --- | --- | --- |
|  | **Dementia cases** | **Sample** | **Hazard Ratio**  **(95% CI)** | **Incidence Rate**  **(95% CI)^b^** | **Dementia cases** | **Sample** | **Hazard Ratio**  **(95% CI)^a^** | **Incidence Rate**  **(95% CI)^b^** |
| **Age** (*p-value* for interaction = 0.10) | | | | | | | | |
| <65 years | 940 | 70,488 | 1 (reference) | 1.10 (1.03-1.17) | 1,073 | 45,436 | 1.77 (1.62-1.93) | 2.00 (1.88-2.12) |
| ≥65 years | 1,699 | 47,271 | 1 (reference) | 3.03 (2.89-3.18) | 2,470 | 43,765 | 1.61 (1.52-1.72) | 4.93 (4.74-5.13) |
| **Sex** (*p-value* for interaction = 0.36) | | | | | | | | |
| Men | 1,408 | 55,956 | 1 (reference) | 2.12 (2.01-2.23) | 1,816 | 42,022 | 1.59 (1.49-1.71) | 3.79 (3.62-3.97) |
| Women | 1,231 | 61,803 | 1 (reference) | 1.64 (1.55-1.74) | 1,727 | 47,179 | 1.67 (1.55-1.80) | 3.09 (2.94-3.24) |
| **Education** (*p-value* for interaction = 0.94) | | | | | | | | |
| College or higher | 977 | 52,629 | 1 (reference) | 1.54 (1.45-1.64) | 1,105 | 34,237 | 1.64 (1.51-1.79) | 2.75 (2.59-2.91) |
| Secondary school  or vocational | 796 | 36,834 | 1 (reference) | 1.80 (1.68-1.93) | 991 | 26,845 | 1.61 (1.46-1.77) | 3.17 (2.98-3.38) |
| No qualification | 866 | 28,296 | 1 (reference) | 2.56 (2.40-2.74) | 1,447 | 28,119 | 1.63 (1.50-1.78) | 4.47 (4.25-4.71) |
| **Socioeconomic status in tertiles** (*p-value* for interaction = 0.24) | | | | | | | | |
| 1 (least deprived) | 581 | 27,304 | 1 (reference) | 1.76 (1.62-1.90) | 596 | 17,554 | 1.49 (1.33-1.67) | 2.87 (2.64-3.10) |
| 2 | 1,554 | 73,038 | 1 (reference) | 1.77 (1.69-1.86) | 2,033 | 53,738 | 1.66 (1.55-1.77) | 3.24 (3.10-3.39) |
| 3 (most deprived) | 504 | 17,417 | 1 (reference) | 2.44 (2.24-2.67) | 914 | 17,909 | 1.68 (1.51-1.88) | 4.51 (4.22-4.81) |

Abbreviations: CI, Confidence Interval.

**^a^**	All models adjusted for age, ethnicity, education, socioeconomic status and APOE-ε4. Specific covariates are dropped from the model where that covariate is the effect modifier of interest.

**^b^**	Incidence rate per 1,000 person-years

**eTable 6 – Probabilities and observed-expected ratios for 41 conditions within 7 clusters in women**

|  |  | **Hypertension, diabetes & CHD** | | **Pain, dyspepsia & depression** | | **Cancer** | | **Thyroid disorders** | | **Pain, osteoporosis & dyspepsia** | | **Asthma & COPD** | | **Pain & hypertension** | |
| --- | --- | --- | --- | --- | --- | --- | --- | --- | --- | --- | --- | --- | --- | --- | --- |
| **Condition** | **Expected** | **P** | **O/E** | **P** | **O/E** | **P** | **O/E** | **P** | **O/E** | **P** | **O/E** | **P** | **O/E** | **P** | **O/E** |
| hypertension | 0.545 | 1.000 | 1.83 | 0.263 | 0.48 | 0.434 | 0.80 | 0.504 | 0.93 | 0.124 | 0.23 | 0.313 | 0.58 | 1.000 | 1.83 |
| depression | 0.109 | 0.091 | 0.83 | 0.231 | 2.12 | 0.071 | 0.65 | 0.071 | 0.65 | 0.120 | 1.10 | 0.088 | 0.81 | 0.028 | 0.25 |
| painful condition | 0.408 | 0.184 | 0.45 | 0.625 | 1.53 | 0.309 | 0.76 | 0.302 | 0.74 | 0.455 | 1.11 | 0.351 | 0.86 | 1.000 | 2.45 |
| asthma | 0.216 | 0.181 | 0.84 | 0.102 | 0.47 | 0.094 | 0.43 | 0.092 | 0.42 | 0.073 | 0.34 | 1.000 | 4.63 | 0.059 | 0.27 |
| coronary heart disease | 0.087 | 0.154 | 1.77 | 0.043 | 0.49 | 0.042 | 0.48 | 0.076 | 0.88 | 0.138 | 1.59 | 0.059 | 0.68 | 0.038 | 0.44 |
| treated dyspepsia | 0.203 | 0.185 | 0.91 | 0.374 | 1.84 | 0.123 | 0.60 | 0.134 | 0.66 | 0.241 | 1.19 | 0.172 | 0.85 | 0.094 | 0.46 |
| diabetes | 0.103 | 0.230 | 2.24 | 0.020 | 0.19 | 0.053 | 0.51 | 0.088 | 0.86 | 0.117 | 1.14 | 0.050 | 0.48 | 0.054 | 0.52 |
| thyroid disorders | 0.207 | 0.078 | 0.38 | 0.117 | 0.57 | 0.119 | 0.58 | 1.000 | 4.83 | 0.086 | 0.42 | 0.135 | 0.65 | 0.000 | 0.00 |
| rheumatoid arthritis | 0.067 | 0.072 | 1.08 | 0.035 | 0.52 | 0.034 | 0.51 | 0.064 | 0.96 | 0.188 | 2.80 | 0.055 | 0.82 | 0.015 | 0.23 |
| COPD | 0.044 | 0.039 | 0.89 | 0.030 | 0.68 | 0.021 | 0.48 | 0.022 | 0.49 | 0.078 | 1.77 | 0.121 | 2.75 | 0.004 | 0.10 |
| anxiety | 0.039 | 0.030 | 0.77 | 0.107 | 2.73 | 0.022 | 0.57 | 0.022 | 0.57 | 0.033 | 0.84 | 0.018 | 0.47 | 0.005 | 0.12 |
| irritable bowel syndrome | 0.062 | 0.037 | 0.59 | 0.191 | 3.08 | 0.031 | 0.49 | 0.025 | 0.41 | 0.039 | 0.62 | 0.049 | 0.79 | 0.012 | 0.20 |
| alcohol problems | 0.001 | 0.001 | 0.76 | 0.004 | 3.91 | 0.001 | 0.72 | 0.000 | 0.00 | 0.002 | 1.68 | 0.001 | 0.90 | 0.000 | 0.00 |
| drug misuse | 0 | 0.000 | 0.00 | 0.001 | 0.00 | 0.000 | 0.00 | 0.000 | 0.00 | 0.000 | 0.00 | 0.000 | 0.00 | 0.000 | 0.00 |
| treated constipation | 0.003 | 0.002 | 0.58 | 0.007 | 2.45 | 0.001 | 0.32 | 0.003 | 1.11 | 0.004 | 1.40 | 0.002 | 0.63 | 0.000 | 0.00 |
| stroke and TIA | 0.042 | 0.082 | 1.94 | 0.016 | 0.37 | 0.023 | 0.55 | 0.029 | 0.70 | 0.074 | 1.76 | 0.015 | 0.36 | 0.016 | 0.39 |
| chronic kidney disease | 0.005 | 0.012 | 2.40 | 0.003 | 0.69 | 0.004 | 0.71 | 0.003 | 0.52 | 0.004 | 0.79 | 0.002 | 0.45 | 0.002 | 0.46 |
| diverticular disease | 0.044 | 0.041 | 0.93 | 0.091 | 2.08 | 0.023 | 0.53 | 0.021 | 0.47 | 0.042 | 0.95 | 0.040 | 0.92 | 0.025 | 0.56 |
| atrial fibrillation | 0.015 | 0.023 | 1.51 | 0.007 | 0.48 | 0.009 | 0.63 | 0.017 | 1.14 | 0.031 | 2.08 | 0.009 | 0.61 | 0.004 | 0.29 |
| peripheral vascular disease | 0.005 | 0.007 | 1.30 | 0.005 | 0.97 | 0.003 | 0.54 | 0.002 | 0.42 | 0.018 | 3.57 | 0.001 | 0.29 | 0.001 | 0.11 |
| heart failure | 0.003 | 0.004 | 1.21 | 0.001 | 0.27 | 0.002 | 0.79 | 0.003 | 0.87 | 0.008 | 2.57 | 0.003 | 0.84 | 0.001 | 0.46 |
| glaucoma | 0.029 | 0.032 | 1.11 | 0.026 | 0.88 | 0.024 | 0.83 | 0.021 | 0.74 | 0.055 | 1.91 | 0.023 | 0.81 | 0.012 | 0.42 |
| epilepsy | 0.012 | 0.010 | 0.84 | 0.008 | 0.64 | 0.010 | 0.86 | 0.011 | 0.92 | 0.039 | 3.29 | 0.009 | 0.72 | 0.000 | 0.03 |
| schizophrenia | 0.006 | 0.005 | 0.87 | 0.005 | 0.86 | 0.002 | 0.27 | 0.010 | 1.64 | 0.012 | 2.00 | 0.004 | 0.62 | 0.000 | 0.00 |
| psoriasis or eczema | 0.056 | 0.042 | 0.75 | 0.089 | 1.58 | 0.027 | 0.47 | 0.029 | 0.52 | 0.092 | 1.64 | 0.087 | 1.55 | 0.015 | 0.26 |
| inflammatory bowel disease | 0.016 | 0.015 | 0.96 | 0.010 | 0.60 | 0.012 | 0.76 | 0.006 | 0.39 | 0.051 | 3.19 | 0.014 | 0.89 | 0.007 | 0.42 |
| migraine | 0.061 | 0.036 | 0.59 | 0.163 | 2.66 | 0.044 | 0.72 | 0.031 | 0.51 | 0.068 | 1.11 | 0.037 | 0.60 | 0.012 | 0.19 |
| chronic sinusitis | 0.013 | 0.007 | 0.56 | 0.031 | 2.35 | 0.009 | 0.68 | 0.006 | 0.46 | 0.012 | 0.94 | 0.020 | 1.53 | 0.002 | 0.13 |
| anorexia or bulimia | 0.001 | 0.001 | 0.76 | 0.001 | 1.39 | 0.001 | 0.81 | 0.000 | 0.21 | 0.003 | 2.50 | 0.001 | 1.40 | 0.000 | 0.00 |
| bronchiectasis | 0.009 | 0.008 | 0.84 | 0.006 | 0.67 | 0.005 | 0.58 | 0.004 | 0.40 | 0.019 | 2.12 | 0.023 | 2.56 | 0.000 | 0.04 |
| Parkinson's disease | 0.004 | 0.003 | 0.75 | 0.001 | 0.28 | 0.003 | 0.66 | 0.001 | 0.34 | 0.014 | 3.57 | 0.004 | 0.91 | 0.000 | 0.00 |
| multiple sclerosis | 0.006 | 0.006 | 1.04 | 0.005 | 0.79 | 0.005 | 0.79 | 0.003 | 0.51 | 0.016 | 2.70 | 0.004 | 0.71 | 0.002 | 0.27 |
| viral hepatitis | 0.004 | 0.002 | 0.54 | 0.009 | 2.31 | 0.003 | 0.70 | 0.004 | 0.94 | 0.003 | 0.65 | 0.002 | 0.60 | 0.001 | 0.20 |
| chronic liver disease | 0.005 | 0.003 | 0.65 | 0.006 | 1.10 | 0.003 | 0.67 | 0.004 | 0.72 | 0.012 | 2.48 | 0.002 | 0.35 | 0.002 | 0.42 |
| osteoporosis | 0.081 | 0.058 | 0.71 | 0.054 | 0.66 | 0.080 | 0.99 | 0.053 | 0.65 | 0.251 | 3.09 | 0.075 | 0.93 | 0.021 | 0.26 |
| chronic fatigue syndrome | 0.001 | 0.006 | 6.09 | 0.026 | 26.02 | 0.005 | 4.80 | 0.006 | 6.36 | 0.005 | 5.47 | 0.010 | 9.56 | 0.000 | 0.41 |
| endometriosis | 0.02 | 0.016 | 0.82 | 0.041 | 2.03 | 0.013 | 0.65 | 0.012 | 0.58 | 0.024 | 1.18 | 0.014 | 0.68 | 0.006 | 0.32 |
| Meniere’s disease | 0.008 | 0.007 | 0.88 | 0.012 | 1.48 | 0.005 | 0.61 | 0.005 | 0.61 | 0.015 | 1.90 | 0.003 | 0.33 | 0.007 | 0.84 |
| pernicious anaemia | 0.011 | 0.008 | 0.69 | 0.005 | 0.50 | 0.002 | 0.21 | 0.026 | 2.34 | 0.027 | 2.50 | 0.005 | 0.50 | 0.003 | 0.30 |
| polycystic ovary | 0.001 | 0.001 | 0.63 | 0.002 | 2.13 | 0.001 | 0.95 | 0.002 | 1.90 | 0.001 | 0.56 | 0.001 | 0.78 | 0.001 | 1.01 |
| cancer | 0.202 | 0.086 | 0.42 | 0.057 | 0.28 | 1.000 | 4.95 | 0.089 | 0.44 | 0.065 | 0.32 | 0.121 | 0.60 | 0.000 | 0.00 |

Abbreviations: O/E, Observed/Expected, P, Probability, CHD, Coronary Heart Disease, COPD, Chronic Obstructive Pulmonary Disease, TIA, Transient Ischemic Attack

Orange cells flag O/E > 1; of these the 3 highest probability conditions > 0.10 are marked as blue.

**eTable 7 – Probabilities and observed-expected ratios for 40 conditions within 6 clusters in men**

|  |  | **Hypertension, pain & dyspepsia** | | **Pain, dyspepsia & prostate disorders** | | **CHD, hypertension & stroke** | | **Asthma, COPD & psoriasis** | | **Diabetes & hypertension** | | **Cancer** | |
| --- | --- | --- | --- | --- | --- | --- | --- | --- | --- | --- | --- | --- | --- |
| **Condition** | **Expected** | **P** | **O/E** | **P** | **O/E** | **P** | **O/E** | **P** | **O/E** | **P** | **O/E** | **P** | **O/E** |
| hypertension | 0.657 | 1.000 | 1.522 | 0.008 | 0.012 | 0.737 | 1.122 | 0.511 | 0.778 | 0.845 | 1.286 | 0.614 | 0.935 |
| depression | 0.07 | 0.071 | 1.014 | 0.142 | 2.029 | 0.055 | 0.786 | 0.055 | 0.786 | 0.033 | 0.471 | 0.041 | 0.586 |
| painful condition | 0.351 | 0.431 | 1.228 | 0.542 | 1.544 | 0.260 | 0.741 | 0.264 | 0.752 | 0.215 | 0.613 | 0.257 | 0.732 |
| asthma | 0.176 | 0.015 | 0.085 | 0.065 | 0.369 | 0.074 | 0.420 | 1.000 | 5.682 | 0.069 | 0.392 | 0.000 | 0.000 |
| coronary heart disease | 0.226 | 0.000 | 0.000 | 0.083 | 0.367 | 1.000 | 4.425 | 0.115 | 0.509 | 0.164 | 0.726 | 0.154 | 0.681 |
| treated dyspepsia | 0.185 | 0.194 | 1.049 | 0.336 | 1.816 | 0.155 | 0.838 | 0.151 | 0.816 | 0.082 | 0.443 | 0.139 | 0.751 |
| diabetes | 0.197 | 0.040 | 0.203 | 0.051 | 0.259 | 0.174 | 0.883 | 0.079 | 0.401 | 1.000 | 5.076 | 0.060 | 0.305 |
| thyroid disorders | 0.047 | 0.049 | 1.043 | 0.075 | 1.596 | 0.044 | 0.936 | 0.031 | 0.660 | 0.043 | 0.915 | 0.033 | 0.702 |
| rheumatoid arthritis | 0.033 | 0.037 | 1.121 | 0.055 | 1.667 | 0.028 | 0.848 | 0.032 | 0.970 | 0.014 | 0.424 | 0.026 | 0.788 |
| COPD | 0.057 | 0.040 | 0.702 | 0.070 | 1.228 | 0.054 | 0.947 | 0.129 | 2.263 | 0.023 | 0.404 | 0.033 | 0.579 |
| anxiety | 0.027 | 0.029 | 1.074 | 0.061 | 2.259 | 0.016 | 0.593 | 0.017 | 0.630 | 0.008 | 0.296 | 0.017 | 0.630 |
| irritable bowel syndrome | 0.022 | 0.024 | 1.091 | 0.057 | 2.591 | 0.009 | 0.409 | 0.017 | 0.773 | 0.004 | 0.182 | 0.012 | 0.545 |
| alcohol problems | 0.004 | 0.004 | 1.000 | 0.008 | 2.000 | 0.002 | 0.500 | 0.003 | 0.750 | 0.000 | 0.000 | 0.003 | 0.750 |
| drug misuse | 0.000 | 0.000 | 0.000 | 0.001 | 0.000 | 0.000 | 0.000 | 0.000 | 0.000 | 0.000 | 0.000 | 0.000 | 0.000 |
| treated constipation | 0.001 | 0.001 | 1.000 | 0.002 | 2.000 | 0.001 | 1.000 | 0.001 | 1.000 | 0.001 | 1.000 | 0.002 | 2.000 |
| stroke and TIA | 0.07 | 0.088 | 1.257 | 0.072 | 1.029 | 0.091 | 1.300 | 0.036 | 0.514 | 0.056 | 0.800 | 0.051 | 0.729 |
| chronic kidney disease | 0.007 | 0.010 | 1.429 | 0.004 | 0.571 | 0.010 | 1.429 | 0.003 | 0.429 | 0.007 | 1.000 | 0.008 | 1.143 |
| diverticular disease | 0.023 | 0.026 | 1.130 | 0.042 | 1.826 | 0.014 | 0.609 | 0.020 | 0.870 | 0.013 | 0.565 | 0.018 | 0.783 |
| atrial fibrillation | 0.034 | 0.042 | 1.235 | 0.054 | 1.588 | 0.028 | 0.824 | 0.018 | 0.529 | 0.018 | 0.529 | 0.028 | 0.824 |
| peripheral vascular disease | 0.007 | 0.007 | 1.000 | 0.010 | 1.429 | 0.011 | 1.571 | 0.004 | 0.571 | 0.007 | 1.000 | 0.004 | 0.571 |
| heart failure | 0.006 | 0.006 | 1.000 | 0.007 | 1.167 | 0.008 | 1.333 | 0.004 | 0.667 | 0.004 | 0.667 | 0.002 | 0.333 |
| prostate disorders | 0.117 | 0.135 | 1.154 | 0.242 | 2.068 | 0.073 | 0.624 | 0.081 | 0.692 | 0.048 | 0.410 | 0.070 | 0.598 |
| glaucoma | 0.035 | 0.040 | 1.143 | 0.056 | 1.600 | 0.024 | 0.686 | 0.025 | 0.714 | 0.025 | 0.714 | 0.030 | 0.857 |
| epilepsy | 0.014 | 0.014 | 1.000 | 0.028 | 2.000 | 0.011 | 0.786 | 0.007 | 0.500 | 0.006 | 0.429 | 0.012 | 0.857 |
| schizophrenia | 0.006 | 0.006 | 1.000 | 0.017 | 2.833 | 0.003 | 0.500 | 0.004 | 0.667 | 0.004 | 0.667 | 0.002 | 0.333 |
| psoriasis or eczema | 0.063 | 0.060 | 0.952 | 0.113 | 1.794 | 0.040 | 0.635 | 0.088 | 1.397 | 0.027 | 0.429 | 0.035 | 0.556 |
| inflammatory bowel disease | 0.016 | 0.014 | 0.875 | 0.030 | 1.875 | 0.011 | 0.688 | 0.015 | 0.938 | 0.010 | 0.625 | 0.013 | 0.813 |
| migraine | 0.022 | 0.025 | 1.136 | 0.058 | 2.636 | 0.008 | 0.364 | 0.015 | 0.682 | 0.002 | 0.091 | 0.015 | 0.682 |
| chronic sinusitis | 0.011 | 0.013 | 1.182 | 0.024 | 2.182 | 0.003 | 0.273 | 0.010 | 0.909 | 0.003 | 0.273 | 0.004 | 0.364 |
| anorexia or bulimia | 0 | 0.000 | 0.000 | 0.000 | 0.000 | 0.000 | 0.000 | 0.000 | 0.000 | 0.000 | 0.000 | 0.000 | 0.000 |
| bronchiectasis | 0.005 | 0.003 | 0.600 | 0.007 | 1.400 | 0.002 | 0.400 | 0.013 | 2.600 | 0.001 | 0.200 | 0.004 | 0.800 |
| Parkinson's disease | 0.006 | 0.006 | 1.000 | 0.014 | 2.333 | 0.005 | 0.833 | 0.003 | 0.500 | 0.002 | 0.333 | 0.007 | 1.167 |
| multiple sclerosis | 0.003 | 0.003 | 1.000 | 0.006 | 2.000 | 0.001 | 0.333 | 0.001 | 0.333 | 0.002 | 0.667 | 0.001 | 0.333 |
| viral hepatitis | 0.004 | 0.003 | 0.750 | 0.011 | 2.750 | 0.002 | 0.500 | 0.002 | 0.500 | 0.001 | 0.250 | 0.005 | 1.250 |
| chronic liver disease | 0.004 | 0.003 | 0.750 | 0.007 | 1.750 | 0.002 | 0.500 | 0.001 | 0.250 | 0.004 | 1.000 | 0.003 | 0.750 |
| osteoporosis | 0.011 | 0.008 | 0.727 | 0.025 | 2.273 | 0.008 | 0.727 | 0.013 | 1.182 | 0.002 | 0.182 | 0.011 | 1.000 |
| chronic fatigue syndrome | 0.003 | 0.003 | 1.000 | 0.009 | 3.000 | 0.002 | 0.667 | 0.002 | 0.667 | 0.002 | 0.667 | 0.002 | 0.667 |
| Meniere’s disease | 0.006 | 0.007 | 1.167 | 0.013 | 2.167 | 0.002 | 0.333 | 0.003 | 0.500 | 0.002 | 0.333 | 0.005 | 0.833 |
| pernicious anaemia | 0.005 | 0.004 | 0.800 | 0.010 | 2.000 | 0.003 | 0.600 | 0.003 | 0.600 | 0.006 | 1.200 | 0.003 | 0.600 |
| cancer | 0.157 | 0.000 | 0.000 | 0.056 | 0.357 | 0.000 | 0.000 | 0.146 | 0.930 | 0.070 | 0.446 | 1.000 | 6.369 |

Abbreviations: O/E, Observed/Expected, P, Probability, COPD, Chronic Obstructive Pulmonary Disease, TIA, Transient Ischemic Attack

Orange cells flag O/E > 1; of these the 3 highest probability conditions > 0.05 are marked as blue.

**eTable 8 - Sex stratified cox-proportional hazards models for the association between multimorbidity clusters and incident dementia in the test sample**

| **Multimorbidity clusters** | **Dementia**  **cases** | **Sample** | **% Participants in cluster** | **Hazard Ratio**  **(95% CI)^a^** | **Incident Rate**  **(95% CI)^b^** |
| --- | --- | --- | --- | --- | --- |
| **Female** |  |  |  |  |  |
| No multimorbidity | 1,231 | 61,803 |  | 1 (reference) | 1.64 (1.55-1.74) |
| Hypertension (100%), pain (50%), dyspepsia (22%) | 81 | 2,156 | 22.9% | 1.56 (1.25-1.96) | 3.11 (2.50-3.87) |
| Cancer (100%) | 48 | 1,599 | 17.0% | 1.42 (1.06-1.90) | 2.59 (1.95-3.44) |
| Asthma (100%), COPD (11%), psoriasis (10%) | 49 | 1,457 | 15.5% | 1.60 (1.20-2.13) | 2.80 (2.12-3.70) |
| Pain (61%), dyspepsia (35%), osteoporosis (16%) | 47 | 1,416 | 15.0% | 1.58 (1.18-2.12) | 2.78 (2.09-3.70) |
| Thyroid disorders (100%) | 28 | 1,062 | 11.3% | 1.16 (0.80-1.69) | 2.21 (1.52-3.20) |
| Hypertension (89%), diabetes (47%), CHD (24%) | 53 | 1,040 | 11.0% | 2.10 (1.59-2.77) | 4.41 (3.37-5.78) |
| Depression (100%), pain (41%), anxiety (14%) | 26 | 704 | 7.5% | 1.92 (1.30-2.83) | 3.07 (2.09-4.51) |
| **Male** |  |  |  |  |  |
| No multimorbidity | 1,408 | 55,956 |  | 1 (reference) | 2.12 (2.01-2.23) |
| Hypertension (100%), CHD (28%), dyspepsia (19%) | 96 | 2,269 | 26.9% | 1.54 (1.26-1.90) | 3.69 (3.02-4.51) |
| Pain (100%), hypertension (69%) | 67 | 2,097 | 24.9% | 1.14 (0.89-1.45) | 2.76 (2.18-3.51) |
| Diabetes (100%), hypertension (84%), CHD (28%) | 88 | 1,401 | 16.6% | 2.28 (1.83-2.83) | 5.79 (4.70-7.13) |
| Asthma (100%), psoriasis (12%), COPD (11%) | 45 | 1,265 | 15.0% | 1.34 (1.00-1.80) | 3.07 (2.29-4.11) |
| Dyspepsia (32%), cancer (26%), CHD (25%) | 42 | 954 | 11.3% | 1.55 (1.14-2.11) | 3.92 (2.90- 5.30) |
| Depression (100%), dyspepsia (21%), anxiety (14%) | 16 | 447 | 5.3% | 1.63 (1.00-2.67) | 3.23 (1.98-5.28) |

Abbreviations: CI, Confidence Interval.

**^a^**	All models adjusted for age, ethnicity, education, socioeconomic status and APOE-ε4

**^b^**	Incidence rate per 1,000 person-years

**eFigure 1a - SABIC values for multimorbidity cluster solutions in females**

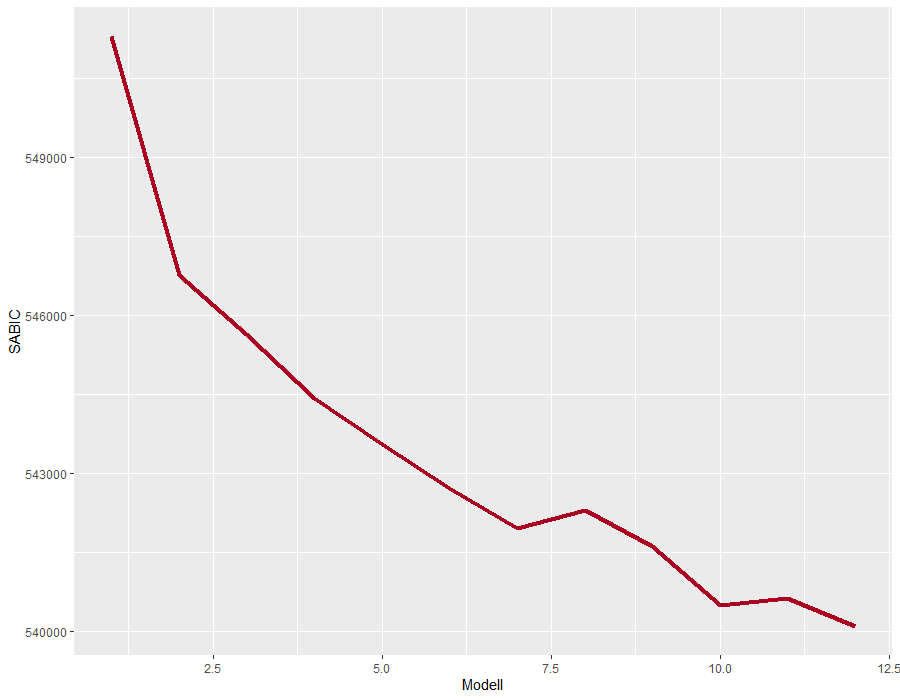

**eFigure 1b - SABIC values for multimorbidity cluster solutions in males**

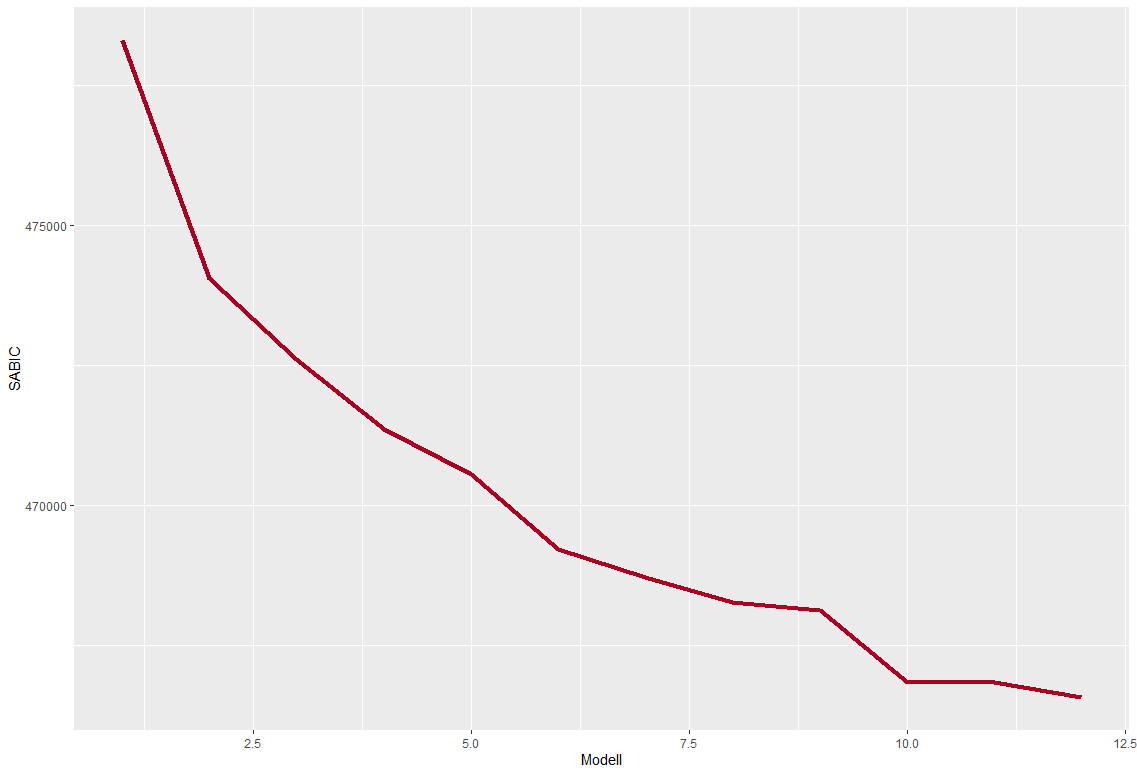
